# Mental health and wellbeing amongst people with informal caring responsibilities across different time points during the COVID-19 pandemic: A population-based propensity score matching analysis

**DOI:** 10.1101/2021.01.21.21250045

**Authors:** Hei Wan Mak, Feifei Bu, Daisy Fancourt

## Abstract

**Aims:** Due to a prolonged period of national and regional lockdown measures during the coronavirus (COVID-19) pandemic, there has been an increase reliance on informal care and a consequent increase in care intensity for informal carers. In light of this, the current study compared the experiences of carers and non-carers on various mental health and wellbeing measures across 5 key time points during the pandemic.

**Methods:** Data analysed were from the UCL COVID -19 Social Study. Our study focused on 5 time points in England: (i) the first national lockdown (March-April 2020; N=12,053); (ii) the beginning of lockdown rules easing (May 2020; N=24,374); (iii) further easing (July 2020; N=21,395); (iv) new COVID-19 restrictions (September 2020; N=4,792); and (v) the three-tier system restrictions (October 2020; N=4,526). We considered 5 mental health and wellbeing measures-depression, anxiety, loneliness, life satisfaction and sense of worthwhile. Propensity score matching were applied for the analyses.

**Results:** We found that informal carers experienced higher levels of depressive symptoms and anxiety than non-carers across all time points. During the first national lockdown, carers also experienced a higher sense of life being worthwhile. No association was found between informal caring responsibilities and levels of loneliness and life satisfaction.

**Conclusion:** Given that carers are an essential national health care support, especially during a pandemic, it is crucial to integrate carers’ needs into healthcare planning and delivery. These results highlight there is a pressing need to provide adequate and targeted mental health support for carers during and following this pandemic.

## Introduction

Prior to the COVID-19 pandemic, there were around 1 in 8 adults (approximately 6.5 million people) providing some form of informal care in the UK, estimated to have a replacement value of £132 billion a year (1). Informal care is defined as unpaid care and support for others (typically family, relatives, friends or neighbours) who may have a disability, chronic illness, mental health problem or other care needs. This can include providing supervision, practical or instrumental care (e.g. shopping, household chores) and personal care (e.g. dressing, bathing, eating, using the bathroom, emotional support) (2,3). With population ageing where the life expectancy for people with long term health conditions has improved, the demand for informal care has increased to meet the needs and to support the sustainability of health and social care system (3). As such, informal care is becoming increasingly important within society.

However, informal care, especially personal care, can be physically and mentally demanding. According to Carers UK (1,4), nearly 1 in 7 of informal carers juggle their caring responsibilities with work, 15% provide over 50 hours of care per week, and 17% care for more than one person. In addition, 3% of the UK general population (approximately more than 1.3 millions people) are ‘sandwich’ carers – people with the dual responsibility of caring for elderly or disabled/sick family members and young children. Often, carers are faced with challenging tasks and stressful situations and are required to maintain high levels of vigilance; this can create chronic stress (5). This is likely to have a negative impact on carers’ personal and social life, and physical and mental health and wellbeing. A substantial literature shows that caring responsibilities have an adverse effect on physical and mental health and health-related behaviours. For instance, it has been shown that people who provide informal care experience higher levels of depression and anxiety, inadequate sleep, higher levels of loneliness, and a higher risk of stroke (1–3,6–8). However, there are also some reported benefits of caregiving, such as self-esteem and sense of meaning (9–11).

During the coronavirus (COVID-19) pandemic, members of the public faced prolonged periods of social distancing, reduced access to local services and community facilities, and restricted face-to-face contacts. Particularly, people considered clinically vulnerable (e.g. older adults aged 70 or above and people with specific medical conditions) faced greatest social restrictions as they were advised to follow stricter advice, often not leaving their homes (“shielding”). For many, this led to an increased reliance on informal care and a consequent increase in care intensity for informal carers (12). Indeed, a report from Carers UK has shown that there were an additional 4.5 million informal carers during 2020 whilst the outbreak of COVID-19 was ongoing (13). Also, limited access to health services means that many carers faced more stressful situations related to care recipients’ medical conditions (14,15). Moreover, to protect those they were caring for, carers themselves had to shield, facing the same tougher restrictions on their social lives and disrupting usual social support networks. There are, consequently, concerns that the mental health of carers was adversely affected during the pandemic. However, whilst there has been wide-spread concern for the negative impact of COVID-19 pandemic on mental health of the public (16–19) and formal carers and other healthcare professionals (20,21), with results suggesting worsening mental health during the pandemic compared with before, less attention has been paid to the mental health and wellbeing of informal carers during the pandemic (13–15,22–25).

Amongst the studies that have been conducted, it has been shown that, since that start of the pandemic, people who providing informal care were likely to be women, younger adults, have children under the age of 18 and have paid work (13). These individuals often experienced a double burden of working or childcare and providing informal care. Some preliminary research has already shown the negative impacts of the pandemic on informal carers. These include increased levels of depression (especially for those who spend 20 hours or more per week on caring) (14), increased mental strain (e.g. the concerns of risk of COVID-19 infection in family (24), increased alcohol consumption and use of illegal drugs (22), increased feelings of frustration (25), and feelings of loss of control and uncertainty (23). However, to date these studies have generally relied on relatively small sample sizes and focused on one time point rather than looking at the evolution of experiences across the pandemic. Further, there has been little research on the impact of informal caring on positive wellbeing during the pandemic.

In light of this, the present study compared the experiences of carers and non-carers on a number of mental health and wellbeing measures, namely depression, anxiety, loneliness, life satisfaction, and a sense that life is worthwhile across various time points during the COVID-19 pandemic. As caring responsibility are socially patterned, with the demographics of carers (e.g. middle-aged adults and females (1)) already linked to less favourable mental health and wellbeing outcomes, this study aimed specifically to disentangle whether the negative impacts of informal caring responsibilities on carers’ mental health and wellbeing were attributable to individual demographics or the role of being an informal carer itself. Whilst direct experimental studies in this context were not feasible or practical, we sought to mimic experimental conditions and to effectively account for the effects of observed confounding factors by using the statistical technique of propensity score matching (PSM).

## Methods

### Participants

This study analysed data from the UK COVID-19 Social Study run by University College London, a longitudinal study that focuses on the psychological and social experiences of adults living in the UK during the COVID-19 pandemic. The study commenced on 21^st^ March 2020 and involves weekly online data collection from participants for the duration of the pandemic. The study is not random and therefore is not representative of the UK population. However, it does contain a heterogeneous sample that was recruited using three primary approaches. First, convenience sampling was used, including promoting the study through existing networks and mailing lists (including large databases of adults who had previously consented to be involved in health research across the UK), print and digital media coverage, and social media. Second, more targeted recruitment was undertaken focusing on (i) individuals from a low-income background, (ii) individuals with no or few educational qualifications, and (iii) individuals who were unemployed. Third, the study was promoted via partnerships with third sector organisations to vulnerable groups, including adults with pre-existing mental health conditions, older adults, carers, and people experiencing domestic violence or abuse. The study was approved by the UCL Research Ethics Committee [12467/005] and all participants gave informed consent. A full protocol for the study is available online at www.COVIDSocialStudy.org.

This study focused on mental health and wellbeing amongst respondents with caring responsibilities across 5 key time points during the pandemic. Given that there were variations in rules and restrictions and the time points that changes to these rules came in across different nations in the UK, we only considered participants who lived in England. We also restricted our sample for each time-point to participants who completed the survey within 7 days of each time point and provided responses to all measures. Participants who opted not to provide details on their demographic background (e.g. gender and household income) were additionally excluded from the analysis. Specifically, our 5 time points were the 5-7 days following the introduction of each of these measures: (i) the first national lockdown (data captured 28 March-04 April 2020; N=12,053); (ii) the beginning of lockdown rules easing (data captured 16-22 May 2020; N=24,374); (iii) further easing (data captured 11-17 July 2020; N=21,395); (iv) new COVID-19 restrictions (data captured 19-25 September 2020; N=4,792); and (v) the three-tier system restrictions (data captured 17-23 October 2020; N=4,526).

### Measures

#### Caring responsibilities

Participants were asked whether they had caring responsibilities for elderly relatives or friends, people with long-term conditions or disabilities, or grandchildren. A binary variable was created to indicate if they had any of the responsibilities.

#### Outcome variables

Five mental health and wellbeing variables were considered. *Depression* was measured using the Patient Health Questionnaire (PHQ-9); a standard instrument for diagnosing depression in primary care which consists of 9 items with responses ranging from “not at all” to “nearly every day”(26). Higher overall scores indicate more depressive symptoms. A*nxiety* was measured using the Generalised Anxiety Disorder assessment (GAD-7); a well-validated tool used to screen and diagnose generalised anxiety disorder in clinical practice and research (27). The assessment includes 7 items with 4-point responses ranging from “not at all” to “nearly every day”, with higher overall scores indicating more symptoms of anxiety. *Loneliness* was measured using the 3-item UCLA-3 loneliness, a short form of the Revised UCLA Loneliness Scale (UCLA-R) (28). Each item is rated with a 4-point rating scale, ranging from “never” to “always”, with higher score indicating greater loneliness. L*ife satisfaction* was measured using the Office for National Statistics (ONS) personal wellbeing question “overall, how satisfied are you with your life nowadays?”; a 10-point scale. S*ense of that life is worthwhile was* measured using the ONS personal wellbeing question “overall, to what extent do you feel the things you do in your life are worthwhile?”; a 10-point scale (29).

#### Covariates

This study considered a set of covariates that could be associated with both caring responsibilities and/or mental health/wellbeing outcomes based on previous empirical research (30,31). These included age groups (age 18-29 vs 30-59 vs 60+), gender (male vs female), ethnicity (white vs ethnic minorities), living arrangement (living alone vs not living alone & not living with children vs not living alone & living with children), marital status (single, never married vs single, divorced/widowed vs in a relationship/married but living apart vs in a relationship/married and cohabiting), education (up to GCSE levels vs A-levels or equivalent vs degree or above), employment status (full-time employment/self-employed vs part-time employment vs economic inactive e.g. students/retires/homemakers/unable to work vs unemployed & seeking work), household income (<£30,000 vs >£30,000 per annum), whether living in an overcrowded household, whether the participant identified as a keyworker, living area (a city vs a large town vs a small town vs a remote area e.g. village/hamlet/isolated dwelling), whether being diagnosed a long-term mental health condition, whether being diagnosed a long-term physical health condition, whether having minor or major stress about COVID-19, whether or not being confirmed or suspected of contracting the COVID-19 virus (confirmed/suspected vs not confirmed/not suspected), perceived social support (measured using an adapted version of the 6-item short form of Perceived Social Support Questionnaire (F-SozU K6); each item is rated on a 5-point scale from “not true at all” to “very true”. Minor adaptations were made to the language in the scale to make it relevant to experiences during COVID-19. Higher scores indicate greater perceived social support; see Supplementary Table 1 for a comparison of changes) (32,33), personality (measured using the Big Five Inventory (BFI-2), which measures 5 domains and 15 facets: neuroticism, extraversion, openness to experience, agreeableness and conscientiousness.

Each item uses a 5-point scale ranging from “strongly disagree” to “strongly agree”. Higher scores indicate greater levels of each domain), and empathy (measured using the Interpersonal Reactivity Index (IRI). Two scales were the focus in the COVID-19 Social Study, empathetic concern/”emotional empathy” and perspective-taking/”cognitive empathy”. Both scales consist of 7 items with a 5-point measure ranging from “does not describe me well” to “describe me very well”, and were standardised. Higher scores indicate greater levels of empathetic concern/perspective-taking).

### Statistics

Our analysis used PSM, a technique that stimulates an experimental setting in an observational data set and creates a treatment group and a control group from the sample (34). One advantage of using PSM over regression approaches is that it controls more effectively for the effects of observed confounders, and hence while results remain observational, bias attributable to confounding can be minimalised significantly. We used PSM to estimate the average treatment effect for the treated (ATT), which is the difference between the average mental health/wellbeing outcomes of participants who had caring responsibilities (carers) and the average outcomes for the same group under the hypothetical scenario that they did not have any caring responsibilities (non-carers).

Specifically, the PSM was performed on an unweighted data, with the kernel matching method with 0.05 bandwidths to perform the matching. Kernel matching uses weighted averages of all individuals in the control group to create the counterfactual outcome, and matches participants in the treatment group to those in the control groups based on the distance of their propensity score. Higher weight is given to the matches whose propensity scores are closer to each other and lower weight to those whose propensity scores are distal from each other (35). A common support condition was imposed to ensure the quality of the matches (31). 95% confidence intervals were computed using bootstrapping techniques with 100 replications. Missing values were handled with list-wise deletion. High quality of matching was achieved; all analyses show Rubin’s B<25%, Rubin’s R of 0.5-2, and a percentage bias of <10% for each covariate (36) (Supplementary Table 2). This suggests that the unobservable heterogeneity reduced significantly after matching. All analyses were carried out using Stata/MP 16.1.

Sensitivity analyses were performed through assigning propensity scores using logistic regression model on a weighted data. Results were very similar but the matching quality was lower. Results are available from the authors upon request.

## Results

### Descriptive statistics

In our complete sample (participants who responded to all measures and who responded in any of the 5 time points), respondents who were informal carers in general were older (age 30-59: 59% vs 47% of non-carers), female (56% vs 50%), not living alone (88% vs 79%), living with a partner (71% vs 64%), had lower educational levels (degree or above: 27% vs 37%), not in full-time employment/self-employment (59% vs 55%), were more likely to be keyworkers (29% vs 20%), not living in a city (72% vs 65%), had a long-term mental health diagnosis (22% vs 19%), and were more likely to be experiencing minor/major stress about COVID-19 (49% vs 42%) (Descriptive statistics were generated from weighted data to show the representative of the demographics of the sample; Table 1).

**Table 1.** Descriptive statistics (UCL COVID-19 Social Study data; weighted)

|  | **Carer**<br>**(N=6,477)** | **Non-carer**<br>**(N=19,977)** |
| --- | --- | --- |
|  | Mean<br>(SE)/% | Mean (SE)/% |
| Age 18-29 | 12.2 | 23.7 |
| Age 30-59 | 59.3 | 47.3 |
| Age 60+ | 28.5 | 29.1 |
| Female | 55.6 | 49.9 |
| Male | 44.4 | 50.1 |
| White | 83.7 | 85.5 |
| Ethnic minority | 16.3 | 14.5 |
| Living alone | 12.0 | 21.5 |
| Not living along & not living with children | 64.9 | 58.4 |
| Not living along & living with children | 23.1 | 20.1 |
| Single, never married | 18.5 | 23.2 |
| Single, divorced or widowed | 10.5 | 12.4 |
| In a relationship/married but living apart | 6.79 | 8.81 |
| In a relationship/married and cohabiting | 64.3 | 55.5 |
| Up to GCSE levels | 37.5 | 30.5 |
| A-levels or equivalent | 36.0 | 32.4 |
| Degree or above | 26.5 | 37.1 |
| Full-time employment/self-employed | 40.6 | 45.1 |
| Part-time employment | 16.2 | 10.9 |
| Economic inactive (e.g. students/retired/homemakers/<br>unable to work) | 41.1 | 41.5 |
| Unemployed & seeking work | 2.13 | 2.49 |
| Household income <£30,000 | 47.3 | 47.1 |
| Household income >£30,000 | 52.7 | 52.9 |
| Living in an overcrowded household | 14.1 | 15.0 |
| Not living in an overcrowded household | 85.9 | 85.0 |
| Keyworker | 28.7 | 19.7 |
| Non-keyworker | 71.4 | 80.3 |
| Living in a city | 28.0 | 35.1 |
| Living in a large town | 20.5 | 19.1 |
| Living in a small town | 27.5 | 24.6 |
| Living in a remote area (e.g. village/hamlet/isolated<br>dwelling) | 24.0 | 21.3 |
| Long-term mental health diagnosis | 22.3 | 19.0 |
| No long-term mental health diagnosis | 77.7 | 81.0 |
| Long-term physical health diagnosis | 48.2 | 41.1 |
| No long-term physical health diagnosis | 51.8 | 58.9 |
| Having minor/major stress about COVID-19 | 49.3 | 42.3 |
| Not having minor/major stress about COVID-19 | 50.7 | 57.7 |
| Confirmed/suspected COVID-19 diagnosis | 11.4 | 10.6 |
| Not confirmed/suspected COVID-19 diagnosis | 88.6 | 89.4 |
| Perceived social support (adapted version of F-SozU K6),<br>ranging from 6-30<sup>1</sup> | 22.0 (0.18) | 22.3 (0.09) |
| Big 5 personality – neuroticism, ranging from 3-21 | 11.5 (0.12) | 11.4 (0.07) |
| Big 5 personality – extraversion, ranging from 3-21 | 12.7 (0.10) | 12.5 (0.06) |
| Big 5 personality – openness to experience, ranging from 3-21 | 14.8 (0.10) | 14.8 (0.05) |
| Big 5 personality – agreeableness, ranging from 3-21 | 15.4 (0.13) | 15.3 (0.05) |
| Big 5 personality – conscientiousness, ranging from 3-21 | 15.9 (0.13) | 15.6 (0.05) |
| Empathy (IRI) – empathetic concern (standardised) | -0.10 (0.03) | -0.17 (0.02) |
| Empathy (IRI) – perspective-taking (standardised) | -0.11 (0.03) | -0.14 (0.02) |
| Depression (PHQ-9), ranging from 0-27<sup>1</sup> | 6.76 (0.26) | 5.84 (0.09) |
| Generalised Anxiety Disorder (GAD-7), ranging from 0-21<sup>1</sup> | 5.22 (0.24) | 4.40 (0.08) |
| Loneliness (UCLA-R), ranging from 3-9<sup>1</sup> | 4.94 (0.07) | 4.92 (0.03) |
| Life satisfaction, ranging from 0-10<sup>1</sup> | 5.92 (0.07) | 6.01 (0.03) |
| Sense of worthwhile, ranging from 0-10<sup>1</sup> | 6.15 (0.06) | 6.09 (0.03) |

Note: <sup>1</sup>The mean and standard error are calculated using the average of the measure across the 5 time-points (i.e. 30 March, Lockdown; 18 May, Easing of lockdown begins; 13 July, Further easing; 21 September, New restrictions; 19 October, Tiered lockdown).

Amongst respondents who provided informal care, when asked to report on the last weekday, 61% reported of not caring for a friend or a relative (suggesting that caring duties were not full-time for the majority of the sample) but around 1 in 5 reported spending 30 mins to 2 hours on informal care, and a further 1 in 5 reported spending three or more hours on informal care (Fig 1).

**Fig 1.**
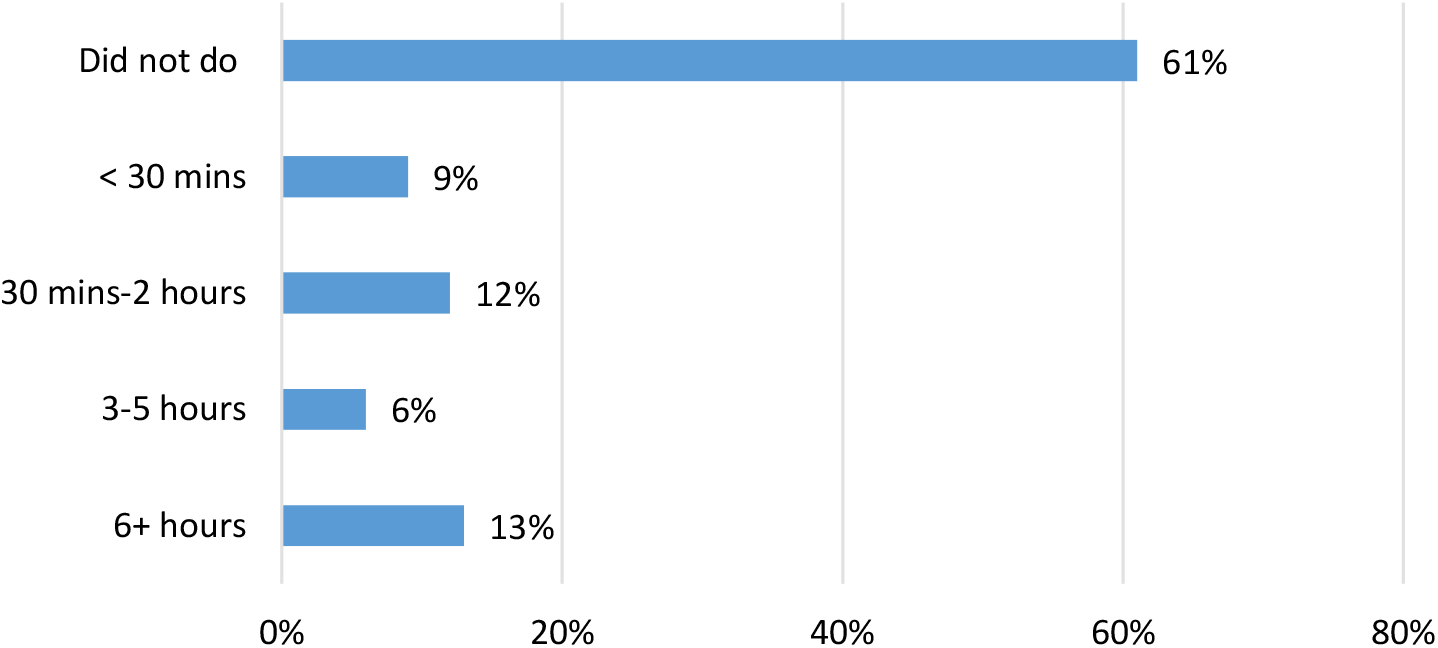
Time spent on caring for a friend or a relative in a day

### Depression (PHQ-9)

Our results show that carers had more depressive symptoms than non-carers across all time-points during the COVID-19 pandemic, with the estimated average treatment effect of being carers on the levels of depression being the strongest during the first national lockdown in March and the most modest during when lockdown measures were most relaxed in July (March: ATT=0.53, 95%CI=0.28, 0.77, p<0.001; May: ATT=0.43, 95%CI=0.27, 0.59, p<0.001; July: ATT=0.39, 95%CI=0.20, 0.59, p<0.001; September: ATT=0.42, 95%CI=0.03, 0.82, p<0.05; October: ATT=0.52, 95%CI=0.18, 0.87, p<0.01) (Fig. 2 & Supplementary Table 2).

**Fig 2.**
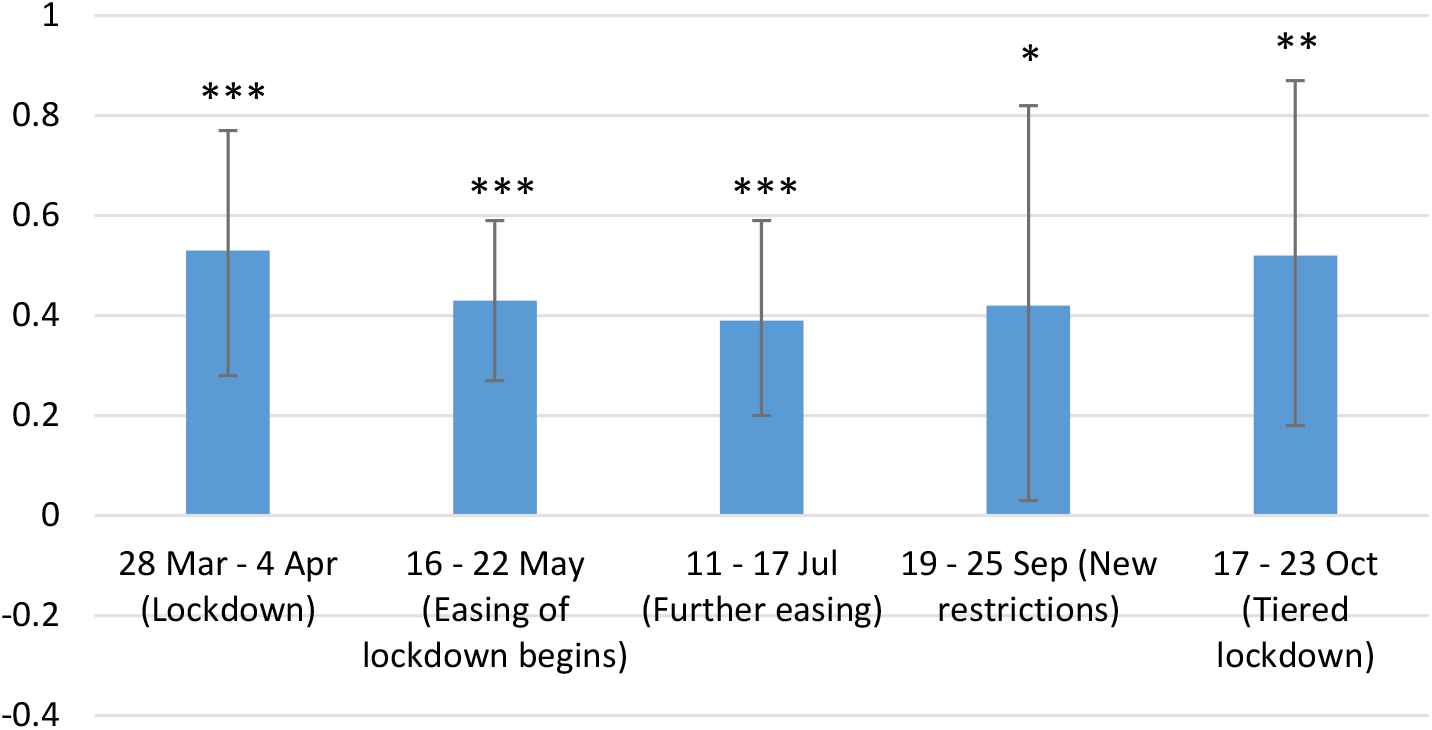
Depression (PHQ-9)

### Generalised Anxiety Disorder (GAD-7)

Similar to depression, we found that caring responsibilities were also associated with higher levels of anxiety across all time-points during the pandemic. The estimated treatment effect of being carers on the anxiety levels were the strongest during the first national lockdown in March and the three-tier system lockdown in October, and were more modest when the restrictions were lower in May and July (March: ATT=0.50, 95%CI=0.28, 0.72, p<0.001; May: ATT=0.38, 95%CI=0.23, 0.52, p<0.001; July: ATT=0.39, 95%CI=0.23, 0.55, p<0.001; September: ATT=0.44, 95%CI=0.11, 0.77, p<0.05; October: ATT=0.49, 95%CI=0.12, 0.85, p<0.01) (Fig. 3 & Supplementary Table 2).

**Fig 3.**
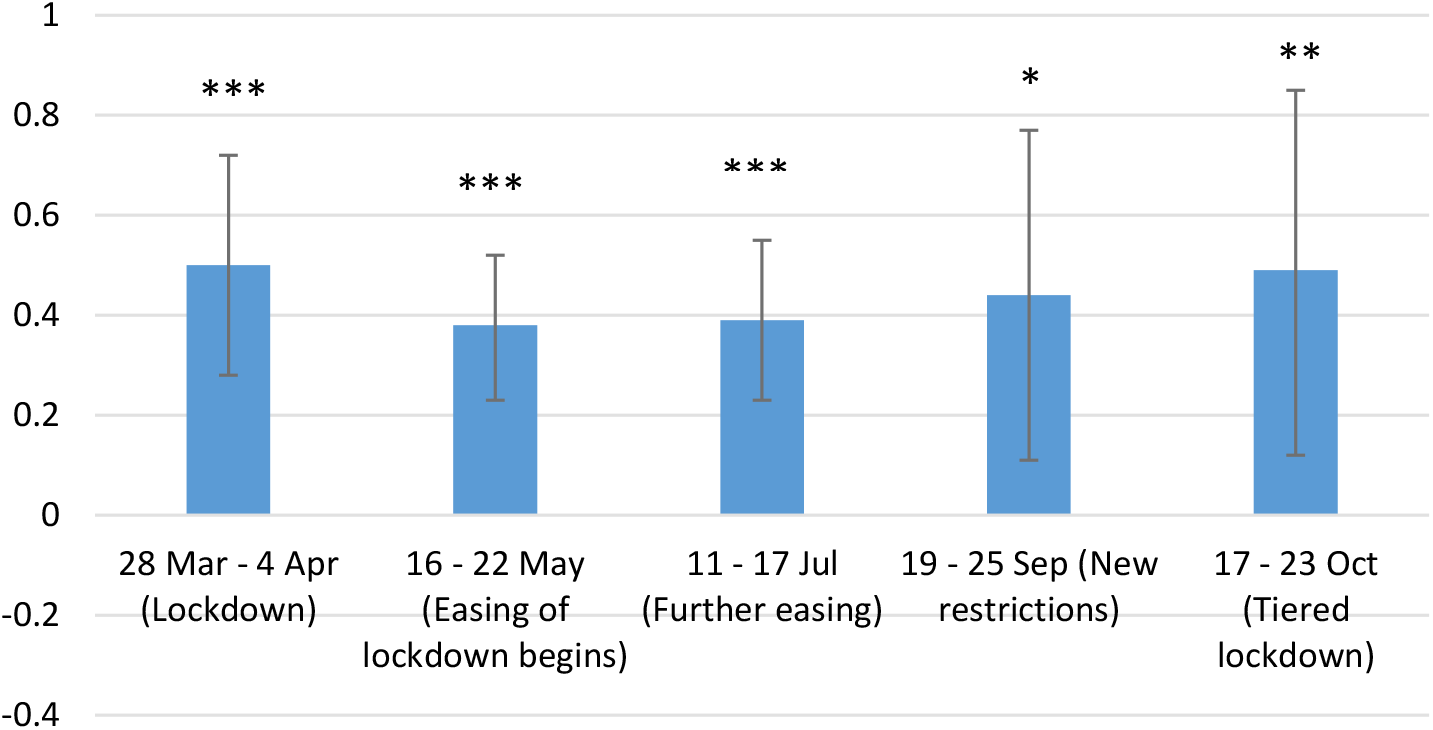
Generalised Anxiety Disorder (GAD-7)

### Loneliness (UCLA-R)

No association was found between caring responsibilities and the levels of loneliness at any of the time points (Fig. 4 & Supplementary Table 2).

**Fig 4.**
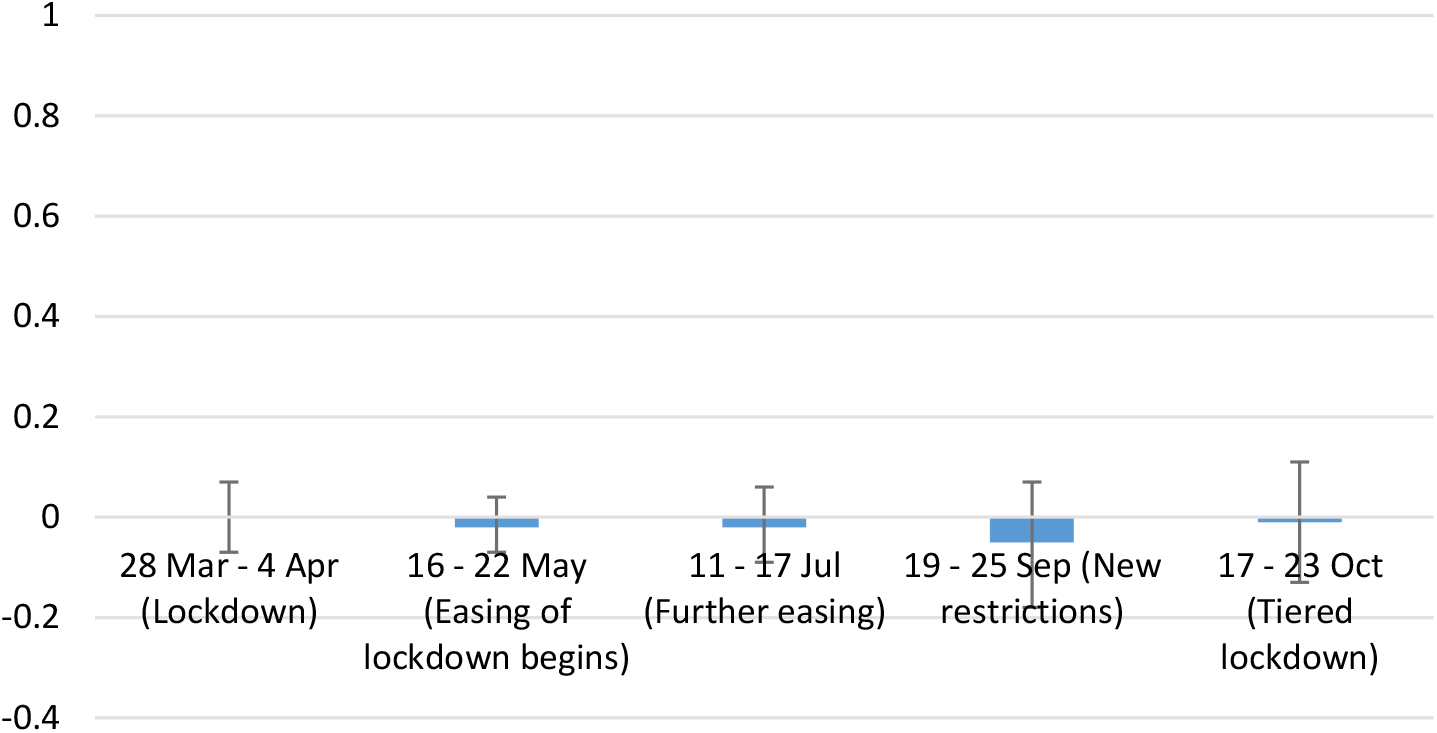
Loneliness (UCLA-R)

### Life satisfaction

No association was found between caring responsibilities and the levels of life satisfaction at any of the time points (Fig. 5 & Supplementary Table 2).

**Fig 5.**
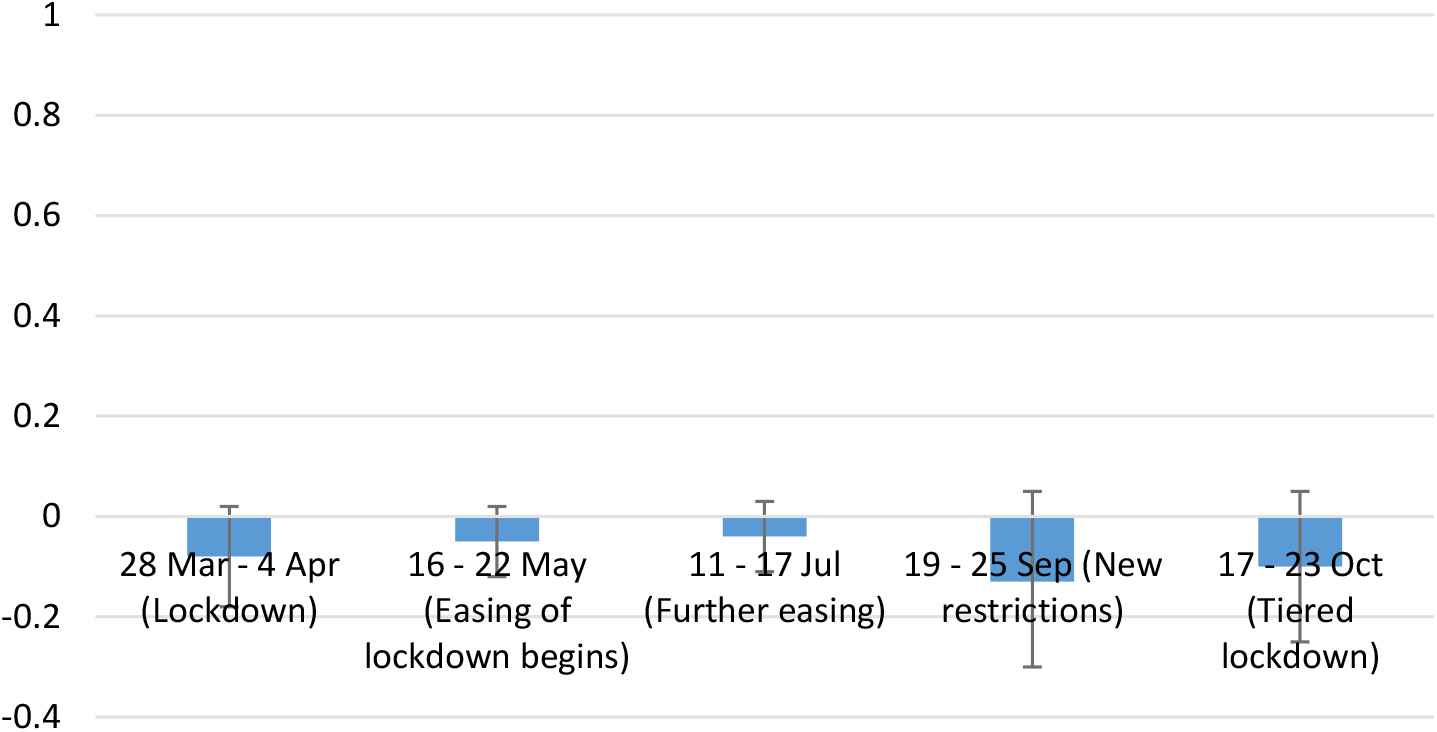
Life satisfaction

### Sense of worthwhileness

Our analysis shows that respondents with caring responsibilities were more likely to have a higher sense of life being worthwhile, but only during the first national lockdown in March (ATT=0.15, 95%CI=0.05, 0.25, p<0.01) (Fig. 6 & Supplementary Table 2).

**Fig 6.**
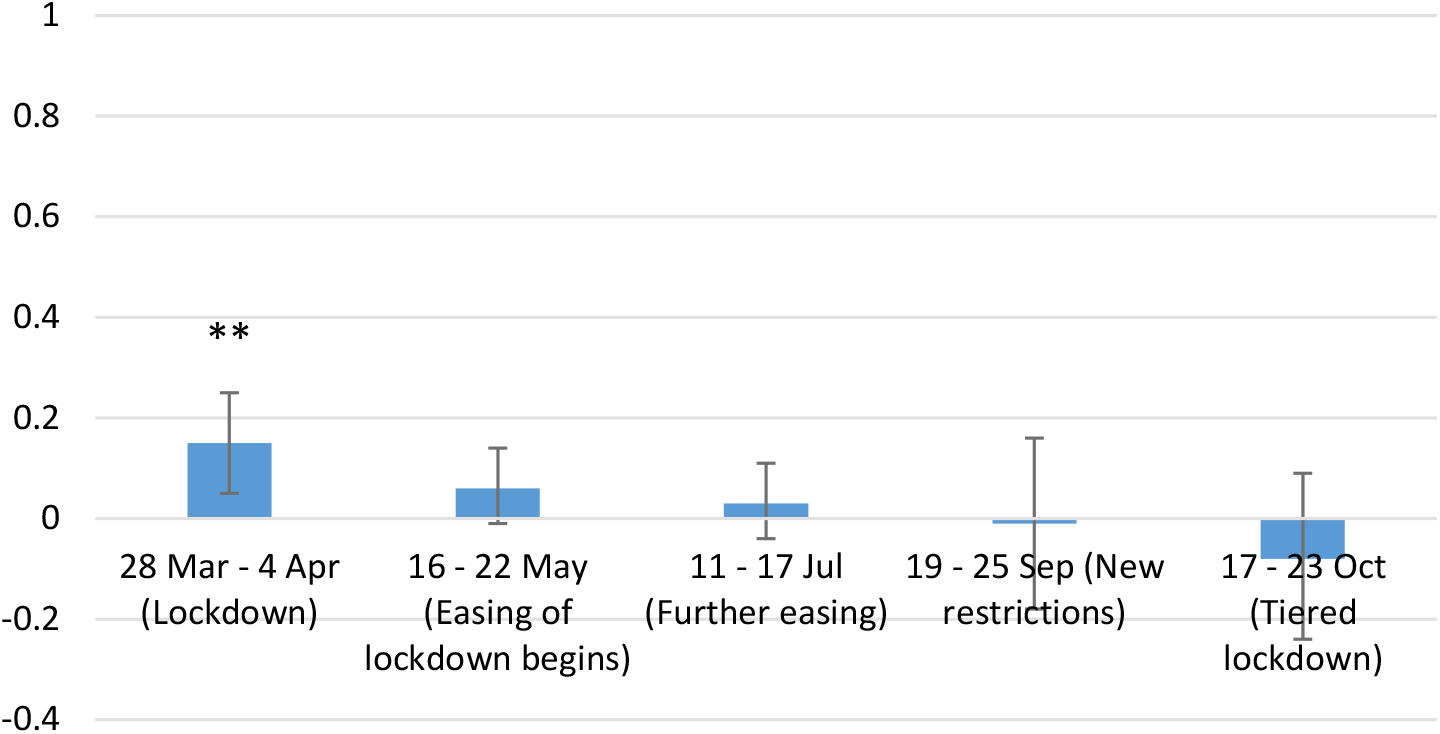
Sense of worthwhile

## Discussion

This study examined the differences in mental health and wellbeing between carers and non-carers across different time points (March to October 2020) during the COVID-19 pandemic using data from England. Results showed that informal carers experienced higher levels of depression and anxiety than people without caring responsibilities consistently across different time points during the lockdown. The relationship between being a carer and poorer mental health was strongest during periods of stricter social restrictions. There was no evidence that carers differed from non-carers in loneliness and life satisfaction. However, we found that carers experienced a greater sense of their lives being worthwhile at the beginning of the first lockdown in England, but no difference was found at later time points when lockdown measures were eased in May and July 2020 or when new restrictions were introduced in September and October 2020.

Our findings that carers had higher levels of depression and anxiety during the COVID-19 pandemic are consistent with existing literature before the pandemic highlighting the mental health burden of informal caring (5,7) and with qualitative and small-scale cross-sectional studies during the COVID-19 pandemic (13,14,23,24). The negative effects of caregiving is commonly explained by the chronic stress model. Care provision creates physical and psychological strain over extended periods of time, which is accompanied by high levels of unpredictability and uncontrollability, frequently requires high levels of vigilance, and creates secondary stress due to competing demands in other roles (5). Chronic stress can lead to psychosocial distress and worsening mental health. The negative experiences associated with caregiving were likely intensified during the COVID-19 pandemic as a result of cuts to formal care, reduced paid working hours, reduced informal support from other relatives or friends, restricted access to health care services, and fear of virus infection (12,14,15,25). Whilst these results, therefore, are not especially surprising, they are still of particular concern in the context of the pandemic. Many vulnerable individuals have been more reliant than ever before on their informal carers. So if poor mental health leads to carer burnout, either affecting care during the pandemic or the willingness and capacity to provide care in the aftermath of the pandemic, this could have substantial implications for those individuals but also for the wider health and social care sector, leaving more work to be carried out by formal carers. In light of this, it is critical that informal carers are provided with adequate targeted mental health support so that they are psychologically able to continue their caring responsibilities.

Aside from the negative psychological impacts, our study has shown that carers may also have experienced a greater sense of life being worthwhile compared to non-carers in the early part of the pandemic. This is in line with previous studies that show the positive experience of caregiving, such as gratification, companionship, meaning, sense of purpose, personal growth and so forth (9–11). Our findings on worthwhileness provide empirical support for the view that both negative and positive experiences may emerge as independent dimensions as a result of caregiving (37). However, it is important to note that the difference in worthwhileness between carers and non-carers was only significant at the beginning of the lockdown. A potential explanation is that as the difficult situation unfolded, the initial greater sense of worthwhileness and appreciation by those they were caring for and others within communities may have been gradually eroded by the stresses of providing that care but also by the decreasing social recognition of the roles carers were playing during the COVID-19 pandemic. Similar patterns have been noted for formal carers, who experienced greater societal appreciation in the early part of the pandemic (including with the national “clap for carers”) but who simultaneously reported decreasing appreciation from the government as the pandemic continued contributing to poorer morale (38).

It is also notable that we found no evidence that carers differs from non-carers in other wellbeing measures, namely loneliness and life satisfaction, which seems to contradict to previous studies that show the correlation between being an informal carer and higher levels of loneliness and lower levels of life satisfaction (although results on life satisfaction are less conclusive as it varies across the types of care, the health conditions of the care recipients, the length of care, etc.) (15,37,39,40). Previous studies have suggested that the reasons for these higher levels of loneliness and lower levels of wellbeing are that care provision is a time and energy consuming task that can restrict carers’ personal and social life. Indeed, a report from Carers UK showed that nearly half of the carers reported not having time to spend on social activities and difficulties being able to leave the house (40). However, such feelings may have changed in the context of the COVID-19 pandemic, even though reports suggest loneliness and social isolation remained a challenge for many carers (15). Due to the lockdown and social distancing measures, face-to-face social activities were greatly restricted for the whole population. As a consequence, caring responsibilities may have reduced feelings of isolation amongst carers as others experienced some of the same social restrictions that they faced before the pandemic, and carers may have felt less of a sense of missing out. As carers had some exemptions from the “stay at home” orders to visit the people they cared for, they might also have been able to maintain companionship during these difficult times. This is supported by a report showing that 2 in 5 young carers and 1 in 5 young adult carers built a stronger relationship with the person they were caring for during the pandemic (22) and nearly 3 in 5 carers reported being able to keep in touch with family and friends despite the lockdown measures (15). It is also possible that the gap between loneliness and wellbeing levels amongst carers and non-carers was reduced as a number of studies have shown that loneliness and poor wellbeing increased for the general population (19).

This study had several limitations. First of all, the UCL COVID-19 Social Study did not use a random sample, therefore our sample is not representative of the population. However, the study does have a large sample size with wide heterogeneity, including good stratification across all major socio-demographic groups, and analyses were weighted based on population estimates of core demographics, with the weighted data showing good alignment with national population statistics and another large scale nationally representative social survey. But we cannot rule out the possibility that the study inadvertently attracted individuals experiencing more extreme psychological experiences, with subsequent weighting for demographic factors failing to fully compensate for these differences. Secondly, the UCL COVID-19 Social Study did not collect any information before the pandemic. Therefore, we were not able to compare the average treatment effect of being a carer before and during the pandemic. Further work is needed to understand if the pandemic has heightened the mental health risk for carers compared with usual times. Thirdly, this study treated carer status as a binary variable, without further exploring the intensity of caregiving, which has important implications for carers’ mental health and wellbeing. It is unknown whether individuals took on new informal caring responsibilities during the pandemic or withdrew from usual informal caring roles. Therefore, future work is needed to examine the role of care intensity and how fluctuating patterns of care affected mental health (14). Relatedly, while PSM can effectively control for observed confounding factors and can stimulate an experimental study on an observational dataset where an experimental setting is not feasible, it is unable to capture unobserved confounding factors. Therefore, future studies are needed to ascertain how experiences of carers vs non-carers varied depending on the type of care provided, the quality of the relationship between carers and the care recipients, and the health conditions of the care recipients. Finally, our analysis focused on comparisons between carers and non-carers at different time points in the pandemic, using PSM to control for confounding variables. However, this analysis did not show how the trajectories of mental health and wellbeing changed for carers vs non carers, and this topic could be the focus of future research.

## Conclusions

The severe lockdown and social distancing measures implemented to control the spread of Covid-19 led to increasing burden for informal carers. The results of this study support some previous literature suggesting that carers were more likely to experience higher levels of depression and anxiety during the pandemic, as in non-pandemic circumstances. But they build on these findings by quantifying this difference and showing how the trajectories of mental health experiences changed in line with changing social restrictions during Covid-19. Carers were also more likely to feel a higher sense of life being worthwhile compared to non-carers, but this effect was attenuated after the first lockdown. In contrast to the existing studies, we found no differences in loneliness and life satisfaction between carers and non-carers, suggesting either that the companionship provided through caring during lockdown and social solidarity in experiencing social restrictions may have offered some emotional benefits to carers, or that worsening levels of personal and social wellbeing amongst non-carers (as documented in previous studies) closed the gap between the experiences of carers and non-carers. As carers are an important support to the national health care support, it is therefore crucial to integrate their needs into healthcare planning and delivery, especially when the health service is stretched as during this pandemic. While there is some existing support available to carers, the results presented here highlight the importance of ensuring adequate and targeted mental health provision to support carers during and following this pandemic so that they are able to continue their vital work.

## Data Availability

Anonymous data will be made available following the end of the pandemic.

## Ethics approval and consent to participate

Ethical approval for the COVID-19 Social Study was granted by the UCL Ethics Committee. All participants provided fully informed consent. The study is GDPR compliant.

## PPI

The research questions in the UCL COVID-19 Social Study built on patient and public involvement as part of the UKRI MARCH Mental Health Research Network, which focuses on social, cultural and community engagement and mental health. This highlighted priority research questions and measures for this study. Patients and the public were additionally involved in the recruitment of participants to the study and are actively involved in plans for the dissemination of findings from the study.

## Author Contributions

FB conceived the study. HWM conducted the data analyses. HWM, FB and DF wrote the first draft. All authors provided critical revisions. All authors read and approved the submitted manuscript.

## Availability of data and materials

Anonymous data will be made available following the end of the pandemic.

## Acknowledgement

We are very grateful to all participants in the COVID-19 Social Study.

## Supplementary

**Supplementary Table 1:** Comparison of items in the original and revised Perceived Social Support Questionnaire (F-SozU K-6)

|  |  |
| --- | --- |
| Original | Adapted for COVID-19<br>In the past week, I feel... |
| I experience a lot of understanding and security from others | I have experienced a lot of understanding and support from others |
| I know a very close person whose help I can always count on | I have a very close person whose help I can always count on |
| If necessary, I can easily borrow something I might need from neighbours or friends | If necessary, I can easily borrow something I need from neighbours or friends |
| I know several people with whom I like to do things | I have people with whom I can spend time and do things together |
| When I am sick, I can without hesitation ask friends and family to take care of important matters for me | If I get sick, I have friends and family who will take care of me |
| If I am down, I know to whom I can go without hesitation | If I am feeling down, I have people I can talk to without hesitation |

**Supplementary Table 2:** **Propensity score matching estimating the association between caring responsibilities and mental health/wellbeing across 5 different time-points during the COVID-19 pandemic**

|  | Depression<br>(PHQ-9) | Generalised<br>Anxiety<br>Disorder<br>(GAD-7) | Loneliness<br>(UCLA-R) | Life<br>satisfaction | Sense of<br>worthwhile |
| --- | --- | --- | --- | --- | --- |
|  | ATT (95%CI) | ATT<br>(95%CI) | ATT (95%CI) | ATT<br>(95%CI) | ATT (95%CI) |
| **28 March – 04**<br>**April 2020**<br>**(Lockdown)** | 0.53 (0.28,<br>0.77)*** | 0.50 (0.28,<br>0.72)*** | -0.00 (-0.07,<br>0.07) | -0.08 (-<br>0.18, 0.02) | 0.15 (0.05,<br>0.25)** |
| Control group | 9,061 |  |  |  |  |
| Treatment<br>group | 2,992 |  |  |  |  |
| Total N | 12,053 |  |  |  |  |
| Mean bias | 0.5 |  |  |  |  |
| Rubin's B | 3.5 |  |  |  |  |
| Rubin's R | 1.16 |  |  |  |  |
| **16-22 May**<br>**2020**<br>**(Easing of**<br>**lockdown**<br>**begins)** | 0.43 (0.27,<br>0.59)*** | 0.38 (0.23,<br>0.52)*** | -0.02 (-0.07,<br>0.04) | -0.05 (-<br>0.12, 0.02) | 0.06 (-0.01,<br>0.14) |
| Control group | 18,417 |  |  |  |  |
| Treatment<br>group | 5,957 |  |  |  |  |
| Total N | 24,374 |  |  |  |  |
| Mean bias | 0.4 |  |  |  |  |
| Rubin's B | 2.8 |  |  |  |  |
| Rubin's R | 1.14 |  |  |  |  |
| **11-17 July 2020**<br>**(Further easing)** | 0.39 (0.20,<br>0.59)*** | 0.39 (0.23,<br>0.55)*** | -0.02 (-0.09,<br>0.06) | -0.04 (-<br>0.11, 0.03) | 0.03 (-0.04,<br>0.11) |
| Control group | 16,141 |  |  |  |  |
| Treatment<br>group | 5,254 |  |  |  |  |
| Total N | 21,395 |  |  |  |  |
| Mean bias | 0.4 |  |  |  |  |
| Rubin's B | 2.8 |  |  |  |  |
| Rubin's R | 1.15 |  |  |  |  |
| **19-25 Sep 2020**<br>**(New**<br>**restrictions)** | 0.42 (0.03,<br>0.82)* | 0.44 (0.11,<br>0.77)* | -0.05 (-0.18,<br>0.07) | -0.13 (-<br>0.30, 0.05) | -0.01 (-0.18,<br>0.16) |
| Control group | 3,571 |  |  |  |  |
| Treatment<br>group | 1,221 |  |  |  |  |
| Total N | 4,792 |  |  |  |  |
| Mean bias | 0.6 |  |  |  |  |
| Rubin's B | 4.5 |  |  |  |  |
| Rubin's R | 1.25 |  |  |  |  |
| **17-23 Oct 2020**<br>**(Tiered**<br>**lockdown)** | 0.52 (0.18,<br>0.87)** | 0.49 (0.12,<br>0.85)** | -0.01 (-0.13,<br>0.11) | -0.10 (-<br>0.25, 0.05) | -0.08 (-0.24,<br>0.09) |
| Control group | 3,362 |  |  |  |  |
| Treatment<br>group | 1,164 |  |  |  |  |
| Total N | 4,526 |  |  |  |  |
| Mean bias | 0.8 |  |  |  |  |
| Rubin's B | 6.0 |  |  |  |  |
| Rubin's R | 1.04 |  |  |  |  |

## References

1. Carers UK. Facts and figures -Carers UK. 2019.

2. Hiel L, Beenackers MA, Renders CM, Robroek SJW, Burdorf A, Croezen S. Providing personal informal care to older European adults: should we care about the caregivers’ health? Prev Med (Baltim). 2015 Jan;70:64–8.

3. NHS. NHS commissioning?» Who is considered a carer? [Internet]. [cited 2020 Dec 9]. Available from: https://www.england.nhs.uk/commissioning/comm-carers/carers/

4. Juggling work and unpaid care: a growing issue [Internet]. London; 2019 Jan [cited 2021 Jan 13]. Available from: http://www.carersuk.org/images/News_and_campaigns/Juggling_work_and_unpaid_care_report_final_0119_WEB.pdf

5. Schulz R, Sherwood PR. Physical and mental health effects of family caregiving. J Soc Work Educ. 2008;44(SUPPL. 3):105–13.

6. Cooper C, Balamurali TBS, Livingston G. A systematic review of the prevalence and covariates of anxiety in caregivers of people with dementia. Int Psychogeriatrics. 2007;19(2):175–95.

7. Pinquart M, Sörensen S. Differences between caregivers and noncaregivers in psychological health and physical health: a meta-analysis. Psychol Aging. 2003;18(2):250–67.

8. Schulz R, Newsom J, Mittelmark M, Burton L, Hirsch C, Jackson S. Health effects of caregiving: the caregiver health effects study: An ancillary study of the cardiovascular health study. Ann Behav Med. 1997 Jun;19(2):110–6.

9. Quinn C, Clare L, Woods RT. The impact of motivations and meanings on the wellbeing of caregivers of people with dementia: a systematic review. C Int Psychogeriatr Assoc. 2010;22(1):43–55.

10. Cohen CA, Colantonio A, Vernich L. Positive aspects of caregiving: rounding out the caregiver experience. Int J Geriatr Psychiatry. 2002 Feb;17(2):184–8.

11. Yu DSF, Cheng S-T, Wang J. Unravelling positive aspects of caregiving in dementia: an integrative review of research literature. Int J Nurs Stud. 2018 Mar;79:1–26.

12. Giebel C, Cannon J, Hanna K, Butchard S, Eley R, Gaughan A, et al. Impact of COVID-19 related social support service closures on people with dementia and unpaid carers: a qualitative study. Aging Ment Health [Internet]. 2020 Sep 21 [cited 2021 Jan 15];1–8. Available from: https://www.tandfonline.com/doi/full/10.1080/13607863.2020.1822292

13. Carers Week 2020 Research Report: The rise in the number of unpaid carers during the coronavirus (COVID-19) outbreak. 2020.

14. Rodrigues R, Simmons C, Schmidt AE, Steiber N. Care in times of COVID-19: The impact of the pandemic on informal caregiving in Austria. SocArXiv. 2020;

15. Caring behind closed doors Forgotten families in the coronavirus outbreak [Internet]. London; 2020 [cited 2021 Jan 14]. Available from: https://www.carersuk.org/images/News_and_campaigns/Behind_Closed_Doors_2020/Caring_behind_closed_doors_April20_pages_web_final.pdf

16. Holmes EA, O’Connor RC, Perry VH, Tracey I, Wessely S, Arseneault L, et al. Multidisciplinary research priorities for the COVID-19 pandemic: a call for action for mental health science [Internet]. Vol. 7, The Lancet Psychiatry. Elsevier Ltd; 2020 [cited 2020 Oct 15]. p. 547–60. Available from: www.thelancet.com/psychiatry

17. Varga T V., Bu F, Dissing AS, Elsenburg LK, Bustamante JJH, Matta J, et al. Loneliness, worries, anxiety, and precautionary behaviours in response to the COVID-19 pandemic: a longitudinal analysis of 200,000 Western and Northern Europeans. Lancet Reg Heal -Eur. 2021 Jan 2;100020.

18. van Tilburg TG, Steinmetz S, Stolte E, van der Roest H, de Vries DH. Loneliness and mental health during the COVID-19 Pandemic: a study among Dutch older adults. Carr D, editor. Journals Gerontol Ser B. 2020 Aug;

19. Pierce M, Hope H, Ford T, Hatch S, Hotopf M, John A, et al. Mental health before and during the COVID-19 pandemic: a longitudinal probability sample survey of the UK population. The Lancet Psychiatry. 2020 Oct 1;7(10):883–92.

20. Shreffler J, Petrey J, Huecker M.The Impact of COVID-19 on Healthcare Worker Wellness: a Scoping Review. West J Emerg Med [Internet]. 2020 [cited 2021 Jan 15];21(5). Available from: http://escholarship.org/uc/uciem_westjem

21. Shaukat N, Ali DM, Razzak J. Physical and mental health impacts of COVID-19 on healthcare workers: A scoping review. Int J Emerg Med [Internet]. 2020 Jul 20 [cited 2021 Jan 15];13(1):40. Available from: https://intjem.biomedcentral.com/articles/10.1186/s12245-020-00299-5

22. My future, my feelings, my family [Internet]. 2020 [cited 2021 Jan 14]. Available from: https://carers.org/resources/all-resources/108-my-future-my-feelings-my-family

23. Giebel C, Cannon J, Hanna K, Butchard S, Eley R, Gaughan A, et al. Impact of COVID-19 related social support service closures on people with dementia and unpaid carers: a qualitative study. Aging Ment Health [Internet]. 2020 Sep 21 [cited 2020 Dec 9];1–8. Available from: https://www.tandfonline.com/doi/full/10.1080/13607863.2020.1822292

24. Chan EYY, Lo ESK, Huang Z, Kim JH, Hung H, Hung KKC, et al. Characteristics and well-being of urban informal home care providers during COVID-19 pandemic: a population-based study. BMJ Open [Internet]. 2020 Nov 17 [cited 2021 Jan 14];10(11):41191. Available from: http://bmjopen.bmj.com/

25. Savla J, Roberto KA, Blieszner R, McCann BR, Hoyt E, Knight AL. Dementia caregiving during the “stay-at-home” phase of COVID-19 pandemic. Carr DS, editor. Journals Gerontol Ser B [Internet]. 2020 Aug 22 [cited 2021 Jan 15];XX:1–5. Available from: https://academic.oup.com/psychsocgerontology/advance-article/doi/10.1093/geronb/gbaa129/5895926

26. Kroenke K, Spitzer RL. The PHQ-9: A new depression diagnostic and severity measure. Psychiatr Ann [Internet]. 2002 [cited 2020 Nov 25];32(9):1–7. Available from: http://jacobimed.org/public/Ambulatory_files/mlove/CurriculumWomenandGeri/Depression/Depre ssion articles/PHQ-9ReviewKroenke.pdf

27. Spitzer RL, Kroenke K, Williams JBW, Löwe B. A brief measure for assessing generalized anxiety disorder: The GAD-7. Arch Intern Med. 2006 May 22;166(10):1092–7.

28. Russell D, Peplau LA, Cutrona CE. The revised UCLA Loneliness Scale: concurrent and discriminant validity evidence. J Pers Soc Psychol [Internet]. 1980 [cited 2020 Oct 12];39(3):472–80. Available from: https://pubmed.ncbi.nlm.nih.gov/7431205/

29. Personal well-being in the UK QMI -Office for National Statistics [Internet]. [cited 2020 Dec 10]. Available from: https://www.ons.gov.uk/peoplepopulationandcommunity/wellbeing/methodologies/personalwellbeingintheukqmi

30. Brookhart MA, Schneeweiss S, Rothman KJ, Glynn RJ, Avorn J, Stürmer T. Variable selection for propensity score models. Am J Epidemiol. 2006;163(12):1149–56.

31. Caliendo M, Kopeinig S. Some practical guidance for the implementation of propensity score matching. DIW Discuss Pap. 2008;485(1):1–29.

32. Kliem S, Mößle T, Rehbein F, Hellmann DF, Zenger M, Brähler E. A brief form of the Perceived Social Support Questionnaire (F-SozU) was developed, validated, and standardized. J Clin Epidemiol. 2015 May 1;68(5):551–62.

33. Lin M, Hirschfeld G, Margraf J. Brief form of the perceived social support questionnaire (F-SozU K-6): Validation, norms, and cross-cultural measurement invariance in the USA, Germany, Russia, and China. Psychol Assess [Internet]. 2019 May 1 [cited 2020 Oct 12];31(5):609–21. Available from: https://pubmed.ncbi.nlm.nih.gov/30589275/

34. Rosenbaum PR, Rubin DB. The central role of the propensity score in observational studies for causal effects. Biometrika [Internet]. 1983;70(1):41–55. Available from: http://www.jstor.org/stable/2335942%5Cnwhttp://about.jstor.org/terms

35. Guo S, Fraser MW. Propensity Score Analysis: statistical methods and applications. USA: Sage Publications, Inc. 2015.

36. Morgan CJ. Reducing bias using propensity score matching. J Nucl Cardiol. 2018;25(2):404–6.

37. Pinquart M, Sörensen S. Associations of caregiver stressors and uplifts with subjective well-being and depressive mood: a meta-analytic comparison. Aging Ment Health. 2004 Sep;8(5):438–49.

38. Borneo A, Dalrymple A, Hadden C, Johnson A, Kiely S, Knape J, et al. Building a better future for nursing: RCN members have their say [Internet]. London; 2020 [cited 2021 Jan 18]. Available from: https://www.rcn.org.uk/professional-development/publications/rcn-builiding-a-better-future-covid-pub-009366

39. Borg C, Hallberg IR. Life satisfaction among informal caregivers in comparison with non-caregivers. Scand J Caring Sci. 2006 Dec;20(4):427–38.

40. Carers UK & Jo Cox Loneliness. The world shrinks: carer loneliness by Carers UK as part of the Jo Cox Loneliness Commission [Internet]. [cited 2020 Dec 9]. Available from: https://www.carersuk.org/for-professionals/policy/policy-library/the-world-shrinks-carer-loneliness-research-report

